# Preoperative functional connectivity by magnetic resonance imaging for refractory neocortical epilepsy

**DOI:** 10.1101/2023.01.10.23284374

**Authors:** Emily A. Johnson, John J. Lee, Carl D. Hacker, Ki Yun Park, Nabi Rustamov, Andy G. S. Daniel, Joshua S. Shimony, Eric C. Leuthardt

**Author notes:** Author Approval: All authors have seen and approved the manuscript. Data Availability Statement: All unidentifiable data, containing no protected health information, de-identified intermediate data, and software used in the present study are available upon reasonable request to the authors.

## Abstract

**Objective:** Patients with refractory epilepsy experience extensive and invasive clinical testing for seizure onset zones treatable by surgical resection. However, surgical resection can fail to provide therapeutic benefit, and patients with neocortical epilepsy have the poorest therapeutic outcomes. This case series studied patients with neocortical epilepsy who were referred for surgical treatment. Prior to surgery, patients volunteered for resting-state functional magnetic resonance imaging (rs-fMRI) in addition to imaging for the clinical standard of care. This work examined the variability of functional connectivity in patients, estimated from rs-fMRI, for associations with surgical outcomes.

**Methods:** This work examined pre-operative structural imaging, pre-operative rs-fMRI, and post-operative structural imaging from seven epilepsy patients. Review of the clinical record provided Engel classifications for surgical outcomes. A novel method assessed pre-operative rs-fMRI from patients using comparative rs-fMRI from a large cohort of healthy control subjects and estimated Gibbs distributions for functional connectivity in patients compared to healthy controls.

**Results:** Three patients had Engel classification Ia, one patient had Engel classification IIa, and three patients had Engel classification IV. Metrics for variability of functional connectivity, including absolute differences of the functional connectivity of each patient from healthy control averages and probabilistic scores for absolute differences, were higher for patients classified as Engel IV, for whom epilepsy surgery provided no meaningful improvement.

**Significance:** This work continues on-going efforts to use rs-fMRI to characterize abnormal functional connectivity in the brain. Preliminary evidence indicates that the topography of variant functional connectivity in epilepsy patients may be clinically relevant for identifying patients unlikely to have favorable outcomes after epilepsy surgery. Widespread topographic variations of functional connectivity also support the hypothesis that epilepsy is a disease of brain resting-state networks.

## Introduction

One third of epilepsy patients will be refractory to medical intervention [1]. Such patients are often evaluated according to protocols that aim to determine whether seizures are focal or generalized in onset. Patients with focal seizure onset zones (SOZs) are typically further evaluated to assess their candidacy for epilepsy surgery, which often provides the best chance for seizure freedom [2].

Months to years of workup are required to characterize, if possible, the borders around SOZs that differentiate epileptogenic from functionally normal brain. Seizure semiology, EEG (scalp and intracranial), and nuclear medicine techniques (MRI, PET, SPECT) remain the mainstays of epilepsy surgery workup [3]. While the overall rate of surgical success (seizure freedom at one year) is approximately 62%, patients with temporal lobe epilepsy have higher surgical success rates than those with neocortical epilepsy [4–9]. Traditional evaluative techniques typically cannot determine well-defined, precisely measured boundaries for localized SOZ for neocortical refractory epilepsy patients, leaving this population with lower likelihoods of surgical success and sometimes without any surgical treatment options.

In recent years, several studies have reported promising findings regarding the use of resting state fMRI (rs-fMRI) for SOZ localization in patients with refractory focal epilepsy. Notably, Boerwinkle et al. investigated SOZs by independent component analysis of rs-fMRI and found that regions exhibiting unique patterns of functional connectivity (FC) overlapped with SOZs determined by intracranial EEG (iEEG) in 90% of cases in a pediatric refractory epilepsy cohort. In light of FC results in that study, several study participants were subsequently offered epilepsy surgery despite previously having been rejected for surgical management due to poor SOZ localization using traditional techniques [10]. Earlier, Stufflebeam highlighted the potential of interictal FC mapping for SOZ localization in five patients by using z scores to compare subject FC to that of 300 normal controls and found that anatomic regions of abnormal FC were closely associated with iEEG determined SOZs [11]. In Boerwinkle and Stufflebeam’s works, clinical outcomes of epilepsy surgeries were not reported, and thus their analyses stopped short of evaluating the relationship between SOZ resections, congruence with regions of abnormal FC, and epilepsy surgery outcomes [10,11]. Siegel et al. compared the rs-fMRI of one post-stroke abulia patient to that of 23 normal controls and found that specific abulia affected networks displayed variant FC despite having normal gray matter metabolism on PET [12]. These findings motivate further investigation into the use of rs-fMRI as a tool for SOZ identification.

This report uses rs-fMRI data to individually compare the pre-operative FC of seven epilepsy surgery candidates to a control cohort. We developed a novel analysis of rs-fMRI inspired by the identification of abnormal FC by Stufflebeam et al. and Siegel et al. [11,12]. Our analysis uses Gibbs distributions for FC by an approach similar to that described previously by Rosa et al. and Gratton et al. [13,14]. The new method directly estimates a map of variations of FC between a single patient and a large collection of FC estimates from 500 healthy normal subjects. We investigated this methodology alongside diagnostic information drawn from the clinical record, semiology, EEG, MRI, PET, SPECT, final decisions for surgical resections, and the Engel classification of surgical outcomes [15].

## Methods

### Clinical Data from Patients

All study protocols were approved by the Institutional Review Board of the Washington University School of Medicine. All patients were evaluated and underwent epilepsy surgery at Barnes Jewish Hospital or Children’s Hospital of Saint Louis. The seven patients included in this case series had refractory neocortical epilepsy, had at least 40 minutes of pre-operative rs-fMRI, were treated with epilepsy surgery, had post-operative structural MRI delineating neurosurgical resection volumes, and had sufficient post-operative clinical data to support Engel classification [15]. Clinical data were collected retrospectively from hospital records from time of initial epilepsy surgery referral through most recent follow-up (MRFU). All patients were diagnosed with refractory neocortical epilepsy with failure of two or more antiepileptic drugs. Age at the time of epilepsy surgery ranged 9 - 59 years (mean 29). Number of years with epilepsy ranged 1.5 - 25.5 years (mean 11.2). SOZs were determined using medical history, seizure semiology, scalp EEG, iEEG, PET/SPECT, and MRI. All patients underwent surgery for epilepsy at the location of their clinically identified SOZs.

Prior to epilepsy surgery workup, patient one had resection of a left temporal ganglioglioma. This patient had typical seizure events both before and after the previous ganglioglioma resection, and ultimately their SOZ was thought to be unrelated. Prior to epilepsy surgery workup, patient two had resection of a right occipital meningioma. Her SOZ was thought to be most likely related to tissue affected by post-surgical change. Following right temporal parietal SOZ resection, patient four had surgical pathology that revealed grade I ganglioglioma. Patient seven had multiple subpial transections but incomplete excision of the SOZ because of proximity to eloquent somatomotor cortex.

Review of post-operative clinical notes through the MRFU determined Engel classifications. Engel class Ia describes patients who are entirely free of seizures following surgery. Engel class IIa describes patients who were initially free of seizures after surgery, but during a period of two years of post-surgical evaluation had rare seizures. Engel class IV refers to those patients with no meaningful improvement in seizure frequency or severity following surgery. Additional details of demographics, comorbidities, semiology, EEG, imaging and operative procedures for all patients are listed in Table 1.

**Table 1.** Clinical, imaging and operative information.

| Patient | 1 | 2 | 3 | 4 | 5 | 6 | 7 |
| --- | --- | --- | --- | --- | --- | --- | --- |
| **Sex (age at surgery)** | F (10-20) | F (50-60) | F (20-30) | M (10-20) | M (20-30) | F (<10) | M (50-60) |
| **Prior surgery & comorbidities** | L Tem ganglioglioma | R Occip meningioma | prior alcohol abuse | ADHD | bipolar disorder, anxiety, Tourette's | perinatal R Par stroke | migraine, HTN, hypothyroidism, T2DM |
| **Type of epilepsy** | CP | CP | CP, GTC | AU, CP | AU, CP | CP, GTC | SP, CP |
| **Semiology** | blinking, L leg automatism, R head and eye deviation | R eye sensory change, L hand automatism | blinking, anger, L head turn then R head turn | vocalization, L arm and leg tonic clonic movement | R head and eye deviation, bimanual automatisms | Eye roll, L arm automatisms, L arm myoclonic jerk | L arm and leg myoclonic jerk, L head and eye deviation, vocalization |
| **Interictal spikes** | L Tem | R ParOccip | R Fron | None | L Tem, L Par | R Par | R Fron |
| **MRI** | prior Res | prior Res | normal | dilated lateral ventricles | normal | R Par atrophy, prior infarct | R Med Fron lesion |
| **PET, SPECT** | none | hypometabolic B Post Fron, Tem, Ant Par & Cer | Hypometabolic B Tem & Cer | hypometabolic R Inf Ant Tem, R Par | hypometabolic B Tem & Cer | hypometabolic B Tem & Cer, hypermetabolic R Par | hypometabolic B Tem, L Tha |
| **Operative summary** | L NeoTem Res | R ParOccip Res | R Fron Res | R TPJ Res<br>R Tem ganglioglioma | L Med Par Res | R Par Res | R Fron SPT |
| **MRFU (years)** | 2.5 | 10 | 3 | 3 | 2 | 9 | 4 |
| **EC at MRFU** | Ia | Ia | Ia | IIa | IV | IV | IV |

Aura (AU), simple partial seizure (SP), complex partial seizure (CP), generalized tonic clonic seizure (GTC); Bilateral (B), Left (L), right (R), frontal (Fron), temporal (Tem), parietal (Par), occipital (Occip), temporoparietal junction (TPJ); Cerebellum (Cer), Thalamus (Tha); Resection (Res), subpial transections (SPT); Parieto-occipital (ParOccip), Neocortical temporal lobe (NeoTem), anterior (Ant), inferior (Inf), medial (Med), posterior (Post); Hypertension (HTN), Type 2 diabetes mellitus (T2DM)

### Healthy Control Subjects

The Brain Genomics Superstruct Project (GSP) Open Access Data Release [16] (https://dataverse.harvard.edu/dataset.xhtml?persistentId=doi:10.7910/DVN/25833) makes publicly available curated and anonymized structural MRI and rs-fMRI from healthy adult control subjects. Pertinently, healthy controls were aged 18-35 years, 42% male, and self-reported to be free of neurological or psychiatric conditions. This work selected the first 500 subjects and released with 744 seconds of rs-fMRI. These first 500 subjects comprised discovery samples for the GSP, separated from additional 500 replication samples.

### MRI Acquisition

For epilepsy patients enrolled at the Washington University School of Medicine, pre-operatively, scanning on the Siemens Trio Tim at 3T acquired rs-fMRI, magnetization-prepared gradient-echo (MP-RAGE) imaging. Rs-fMRI used a gradient echo echo-planar imaging (EPI) sequence with 4 mm isotropic voxel resolution, 2000 ms TR, 27 ms TE, and 90-degree flip angle. Patients were instructed to gaze upon a visual target and rest motionless without sleeping. Post-operatively, structural MRI with fluid-attenuated inversion recovery, T1-weighting, and T1-weighting with gadolinium contrast were acquired.

For healthy controls enrolled in the Genetics Superstruct Project, scanning on matched Siemens Tim Trio scanners at 3T had acquired rs-fMRI and MP-RAGE imaging. Rs-fMRI used an EPI sequence with 3 mm isotropic voxel resolution, 3000 ms TR, 30 ms TE, and 85-degree flip angle. Subjects rested with eyes open for scanning.

### MRI Preprocessing, Co-registration & Segmentation

We preprocessed all rs-fMRI data for analyses of FC using methods previous described [17]. Pertinently, pre-processing included timing corrections for interleaved EPI slices, correction of head motions within and between rs-fMRI scanning series, co-registration of all rs-fMRI frames to a standardized atlas constructed to match the age of patients and the characteristics of the Siemens Trio scanner, isotropic spatial filtering with Gaussian kernels of 6 mm full width at half maximum, Butterworth temporal filtering with passband below 0.1 Hz, and removal of time series regressors from white matter and cerebral spinal fluid for purposes of variance reduction in the time series. Also, the first five rs-fMRI frames of each scanning series were excluded to remove transient precession of magnetization. For healthy controls, rs-fMRI frames were censored using a conventional threshold of 0.5% root-mean-square intensity variations [18]. Consequently, for 500 healthy controls, 130-246 (mean 229, std. dev. 20) time frames from 6.5-12.3 (mean 11.5, std. dev. 1.0) minutes of rs-fMRI were used for analysis. However, for epilepsy patients, conventional thresholds for frame censoring excluded as much as 75% of frames. Given the possibility that epilepsy patients may have rs-fMRI intensity variability arising from pathophysiological processes and that the correlation of such increased intensity variability with spurious head motion is unknown, no rs-fMRI frames from epilepsy patients were censored. Inspection of correlation matrices for epilepsy patients with and without censoring revealed that censoring effects were not uniformly distributed but associated with correlation blocks belonging to resting-state networks (RSNs). For each epilepsy patient, exactly 1200 time frames from 40 min of rs-fMRI were used for analysis.

For each patient all images were co-registered to one another and to standardized atlases using 4dfp (https://4dfp.readthedocs.io) and FSL (https://fsl.fmrib.ox.ac.uk). Pre-operative rs-fMRI frames were temporally averaged to produce an anatomical estimator that included susceptibility-induced warping. These were co-registered using 12 affine degrees of freedom onto pre-operative MP-RAGE images at 1 mm isotropic resolution. Similarly, the most contrastive delineation of surgical resection in post-operative imaging, typically inversion recovery or T1-weighted imaging with gadolinium, were registered using 12 affine degrees of freedom onto pre-operative MP-RAGE images. Pre-operative MP-RAGE images were registered with rigid-body degrees of freedom to a standardized atlas. Resection volumes were determined and segmented using post-operative imaging and semi-automated labeling tools from ITK-SNAP [19]. By composition of affine transformations and numerically inverted affine transformations, rs-fMRI, post-operative imaging, and resection volumes were all co-registered to the standardized atlas. Patient one had prior surgery for left basal temporal ganglioglioma and patient two had prior surgery for right occipital meningioma. These prior, smaller resection volumes were measured using imaging acquired immediately following the tumor surgeries and accounted so as to visualize secondary surgical resections made specifically for epilepsy treatment. Co-registered resection volumes provided visual summaries of the final clinical assessment of SOZs.

We created masks for definition of gray matter using methods previous described [17]. Pertinently, a separate set of 21 subjects of age 23-35 years (mean 27.6) had provided MP-RAGE and rs-fMRI acquired on a Siemens Allegra scanner at 3T. Rs-fMRI used an EPI sequence with 4 mm isotropic voxel resolution, 2160 ms TR, and 90-degree flip angle. MP-RAGE provided parcellations and segmentations, through FreeSurfer (https://surfer.nmr.mgh.harvard.edu), which formed cortical and subcortical gray matter regions. Subject-averaged rs-fMRI with thresholding provided additional masking to remove areas containing large susceptibility distortions. In total, the mask for gray matter enclosed 18,611 voxels in standardized coordinates. Notably, the cerebellum was excluded from the mask.

We also used cortical and subcortical regions of interest (ROIs), based on developments from Power et al., Gordon et al., and Seitzman et al. [20–22], for purposes of visualization. These curated ROIs possessed high correlations within RSNs and reduced correlations between RSNs. The classification of ROIs to RSNs were validated against published characteristics as well as independent testing data. We used the intersection of our gray matter mask with 300 regions defined by Seitzman et al. to generate 201 ROIs confined to gray matter, excluding cerebellum, and excluding susceptibility distortions. The 201 canonical ROIs provided ordered sampling of RSNs: somatomotor dorsal, somatomotor lateral, cingulo-opercular network, auditory, default mode network, parietal memory network, visual, frontoparietal network, salience, dorsal attention network, basal ganglia, thalamus, medial temporal lobe, and reward.

### Analysis of Functional Connectivity

We represented FC using the method of seeds [17]. Specifically, we defined a seed to be a single voxel at position ***x*** of an rs-fMRI time series, following spatial normalization and confined to the gray matter mask. We conventionally assigned FC using the Pearson product moment correlation coefficient for time series between any two seeds, *R*(***x, x***). The complete data object for FC is a mapping of seeds within a three-dimensional manifold to seeds within a dual three-dimensional manifold, but the mapping is conventionally reshaped to map a vector of voxels to a dual vector of voxels, represented as a correlation matrix. For purposes of examining variations of FC in individuals, we computed the expected FC for healthy controls as follows. First, we created correlation matrices for rs-fMRI from each subject, *c*, from the set of 500 healthy control subjects, *C*. Second, we applied Fisher’s z-transformation to map correlations from subject *c* to real-valued z-scores, *z* (*R*_*c*_(***x, x*′**)). Finally, we averaged z-scores over all healthy control subjects, but retained seed positions, to obtain ⟨*z* (*R*_*c*_(***x, x*′**))⟩_*c*∈*C*_,. We defined our metric for variation of FC to be the absolute difference between z-scored correlations from an individual and the averaged z-scored correlations for all healthy controls,

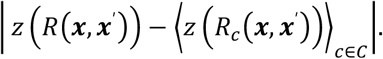

For probabilistic interpretations, we regarded the variation of FC, or absolute difference of z-scored correlations, in the framework of an energy model and Gibbs distribution. As previously detailed by Rosa et al. [13] and implemented by Gratton et al. for the analysis of variations of functional connectivity in Parkinson’s disease [14], a Gibbs distribution can advantageously preserve detailed information contained in data objects such the seeds for RSNs defined in this work. Specifically, a Gibbs distribution enables the entire correlation matrix of size 18,611 × 18,611 for an individual to be a single point in the configuration space of the distribution. By avoiding mass univariate analysis of summary statistics and avoiding adjustments for multiple testing, there is increased statistical power. Formally, we defined a Gibbs distribution

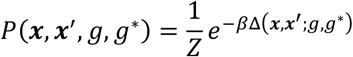

with

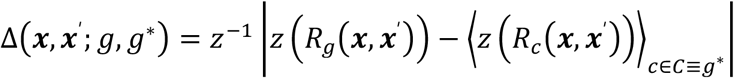

and

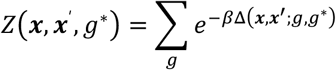

such that *g* indicates the connectome for an individual; *g*∗ indicates the connectome describing centrality; *β* describes the spread of connectomes; Δ incorporates *z*^−1^, the inverse of Fisher’s z-transformation, for the metric of variation of FC; and *Z* is the normalization, which preserves connectome seed positions. We used the average of z-scores for healthy controls to estimate *g*∗. We made no attempt to estimate the spread of connectomes and assigned *β* = 1 for improved numerical precision. Similarly, Δ incorporates *z*^−1^ to bound the metric distance between connectomes to [−11], thereby improving numerical precision. We estimated the normalization, *Z*, by simply computing the sum of exponentials corresponding to the set of 500 healthy control subjects. While providing an empirical adjustment of the normalization at each seed position, our estimate of *Z* cannot provide a true normalization, a computational problem which is intractable in general [23]. For interpretability, we compared the probabilistic divergence of *P*(***x, x***′, *g, g*∗) to its value for *β* = 0, at which the distribution width diverges, centrality is lost, and all parameterizations involving seeds are lost. We denote the latter with *P*_1_. Consequently, the probabilistic divergence, −log(*P*(***x, x***′, *g, g*∗)/*P*_0_), is an adjusted score that describes information. For additional interpretability, we marginalized data objects with respect to the second seed position, using Fisher’s z and its inverse to reduce averaging bias, and obtaining objects denoted −log(*P*(***x***, *g, g*∗)/*P*_0_). For brevity, we omit indicators for connectomes, *g* and *g*∗, hereafter.

For visualizations, and only for visualizations, we additionally resampled 18,611 gray matter voxels using intersections with each of 201 canonical ROIs, described above, so as to create interpretable renderings of correlation matrices. We resampled correlations, applied Fisher’s z-transformation, and then arithmetically averaged z-scores within each canonical ROI. The resulting correlations matrices had size 201 × 201, which could be easily displayed with color representations in figures.

All analyses were computed using Matlab R2019b-R2021a (Mathworks, Natick, MA) and Violin Plots for Matlab (https://github.com/bastibe/Violinplot-Matlab). Project source codes are available (https://github.com/jjleewustledu/mlafc).

## Results

MRFU ranged from 2-10 years with a mean MRFU time of 4.8 years. At MRFU, three patients had Engel Ia, one patient had Engel IIa, and three patients had Engel IV outcomes. All three patients with Engle IV outcomes had seizure recurrence, back to pre-surgical baselines, within one year of epilepsy surgery and remained Engle IV through MRFU.

Patients’ known years with epilepsy at the time of epilepsy surgery ranged from 1.5-25.5 years with a mean of 12.7 years. Patients with Engle Ia and IIa had epilepsy histories that ranged from 1.5-17.5 years. Patients with Engle IV outcomes tended to have longer epilepsy histories at 9, 17.5, and 25.5 years. Despite differences in epilepsy history duration, all patients had localizing scalp EEG and iEEG on pre-operative workup. There were no clearly identifying trends in pre-operative seizure semiology, imaging results, EEG results, or medical histories to distinguish patients with Engle IV outcomes.

Patient five (Engel IV) experienced a gradual increase in seizure frequency following surgery with seizure semiology similar to that of pre-operative events and subsequently underwent a second epilepsy surgery evaluation. Patient six (Engel IV) experienced a return of typical seizure events within several months of surgery, subsequently had treatment with vagal nerve stimulation, and was later noted to have cognitive/executive functioning deficits. Patient seven (Engel IV) began having typical seizures within days of surgery, had infection complications post-operatively that resulted in a one-month hospitalization with reoperation for surgical site drainage, and had new onset chronic mild left hemiparesis. Notably, patient seven was the only patient in our cohort to receive multiple subpial transections as opposed to complete resection of SOZ.

Figures 1 through 3 illustrate the methodology for measuring variations of FC. Figure 1 shows averaged correlations between canonical seeds for 500 healthy control subjects. Highly correlated diagonal blocks within RSNs and patterns of moderate correlations in off-diagonal blocks reproduced previously reported FC for healthy control subjects [14,24]. Figure 2 indicates variations of FC for each epilepsy patient using the absolute difference of correlations between each patient and the control average. Patients classified as Engel Ia had modestly increased variability of FC in somatomotor, default mode, frontoparietal, and salience networks, both diagonal and off-diagonal. Patients classified as Engel IV had the largest variability of FC in multiple RSNs, both diagonal and off-diagonal. Figure 3 indicates variations of FC for each epilepsy patient using an adjusted score for each patient. FC scores greater than zero were wide-spread for Engel IV patients.

**Figure 1.**
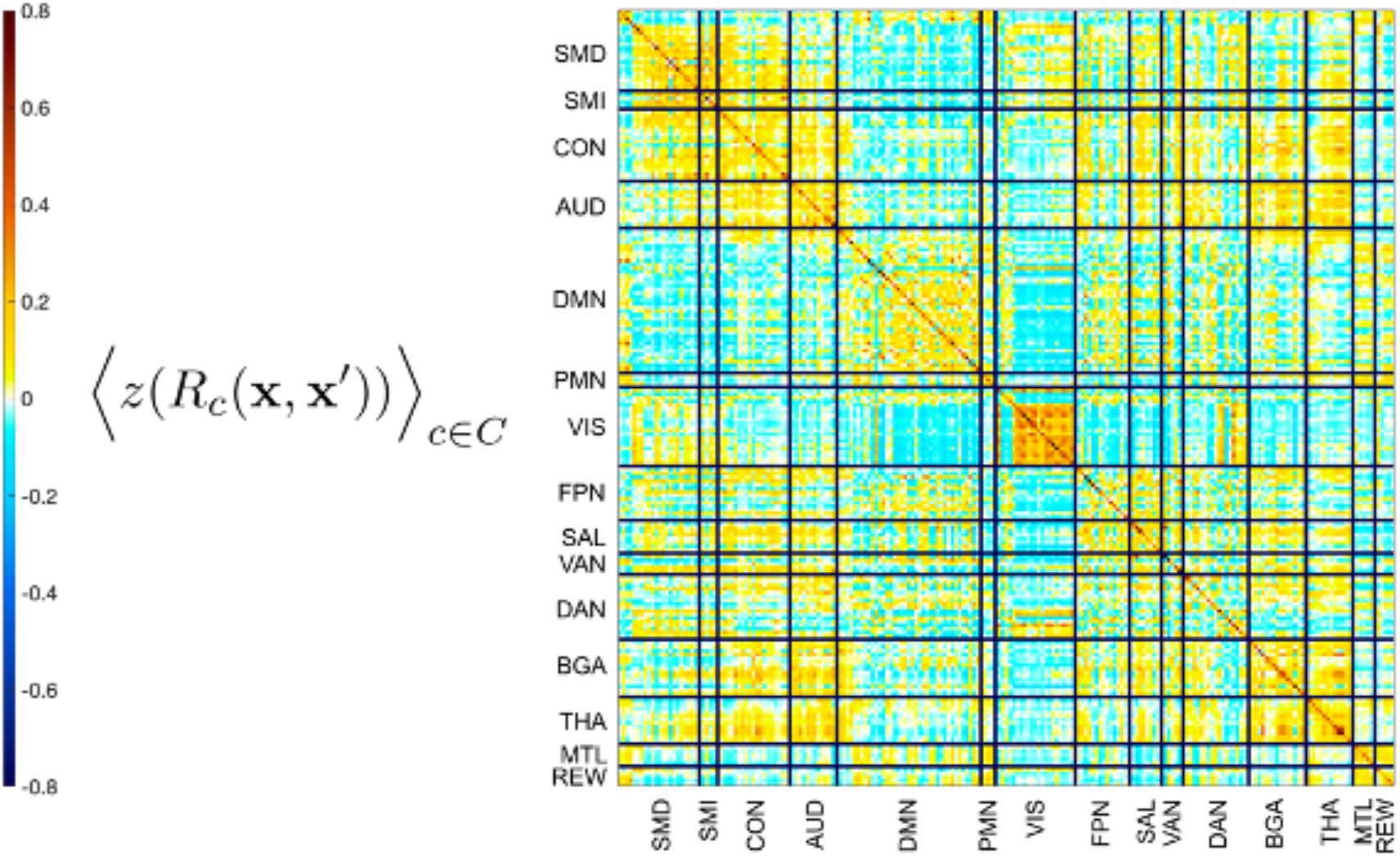
Illustrates the functional connectivity of 500 healthy control subjects, c, drawn from the Genetics Superstruct Project, C. Resting-state fMRI from each control subject formed matrices of Pearson correlations, R_c_, for time series following denoising, spatial smoothing, low-pass temporal filtering, global signal regression, spatial normalization, and sampling from the intersection of masks for gray matter and masks for canonical regions of interest for resting-state networks. **x** and **x**’ denote voxel positions selected by masks. Following conventions for functional connectivity, the Fisher z-transformation, z(), maps correlations to real numbers. Angled brackets denote averaging over all control subjects from the control cohort, c ∈ C. Labels for canonical regions of interest denote: somatomotor dorsal, somatomotor lateral, cingulo-opercular network, auditory, default mode network, parietal memory network, visual, frontoparietal network, salience, dorsal attention network, basal ganglia, thalamus, medial temporal lobe, and reward. Warmer colors in the represented matrix indicate greater positive correlations between canonical regions and cooler colors indicate greater negative correlations. High correlations in the block diagonal elements and patterns of correlations for off-diagonal elements are consistent with previous reports of functional connectivity in healthy controls.

**Figure 2.**
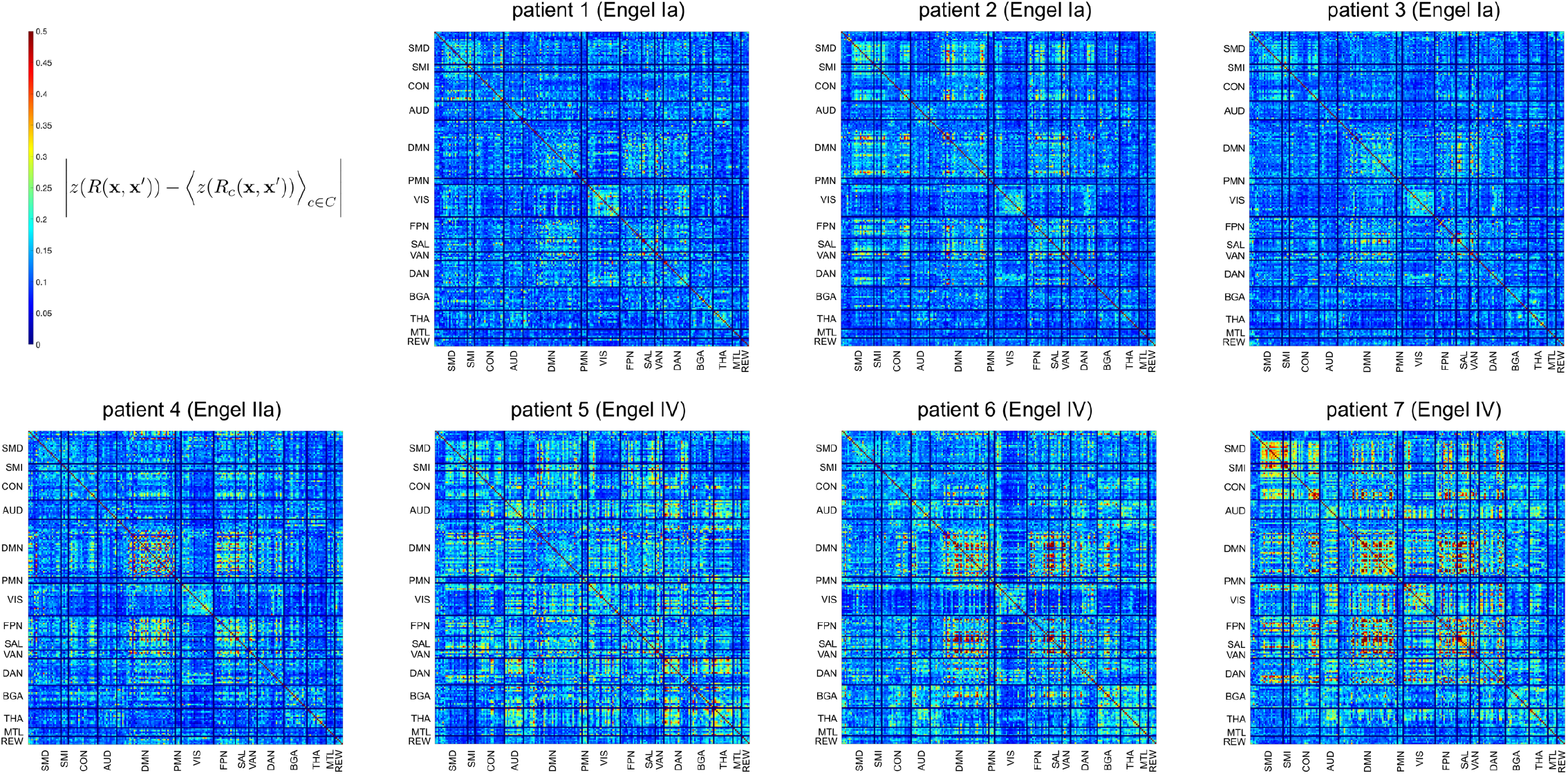
Demonstrates the variability of functional connectivity of epilepsy patients compared to the averaged functional connectivity of 500 healthy control subjects drawn from the Genetics Superstruct Project, which is detailed in Figure 1. Colored representations of matrices describe the absolute difference of functional connectivity computed for each epilepsy patient from the averaged functional connectivity of healthy control subjects. Absolute differences operate on real values yielded by Fisher z-transformed correlations, preserving the domain of canonical regions of interest for resting-state networks, denoted by **x** and **x**’. Warmer colors indicate greater variation of functional connectivity of patients from controls.

**Figure 3.**
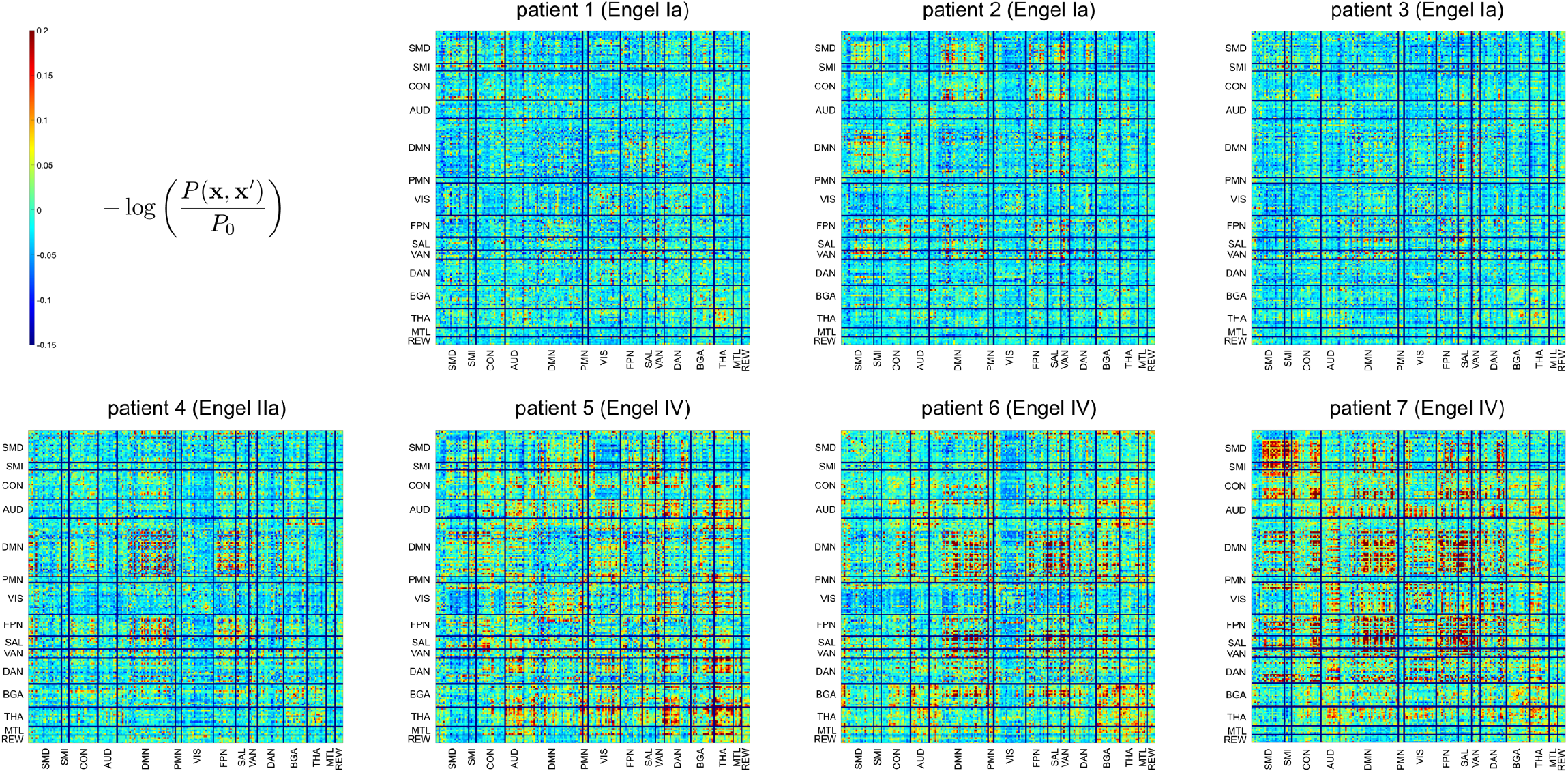
Describes Gibbs distribution, P(**x, x**’), for the absolute differences in functional connectivity between epilepsy patients and the averaged functional connectivity of 500 healthy control subjects drawn from the Genetics Superstruct Project, which is detailed in Figure 2. Compared to the contrast of absolute differences shown in Figure 2, the Gibbs formulation provides an exponential distribution of contrasts and an approximate partition function that normalizes each color-represented matrix element, at canonical regions of interest **x** and **x**’, with functional connectivities from healthy controls. For clarity, we list only deterministic anatomical regions, **x** and **x**’. The Gibbs distribution, however, describes samples drawn from the set of possible functional connectivities for an entire brain, the anatomical regions of which have biologically characteristic correlation structures. The uniform distribution, P_0_, which is the Gibbs distribution for high dispersion, simplifies accounting for the finite sample of 500 healthy control subjects, and enables interpreting each color-represented matrix element of -log(P(**x, x**’)/P_0_) to be an information divergence from the uniform distribution.

Figure 4 shows all patients, their Engel classifications, the anatomical map of scored variants of FC, the co-registered locations of volumes of surgical resection, and the original post-surgical imaging used to segment volumes of resection. Patients classified as Engel IV demonstrated the greatest scores and broadest anatomical distributions for variant FC. Patient five had minimal overlap of variant FC and volume of resection, but variant FC was elevated throughout the brain. Patient seven had high overlap of variant FC and volume of resection but also had generalized elevation of variant FC throughout the neocortex. Patients one through four were Engel Ia or IIa, and had focally variant FC with regional elevations adjacent or within the volume of resection. Notably, the maximally variant FC was outside the resection volume for all patients.

**Figure 4.**
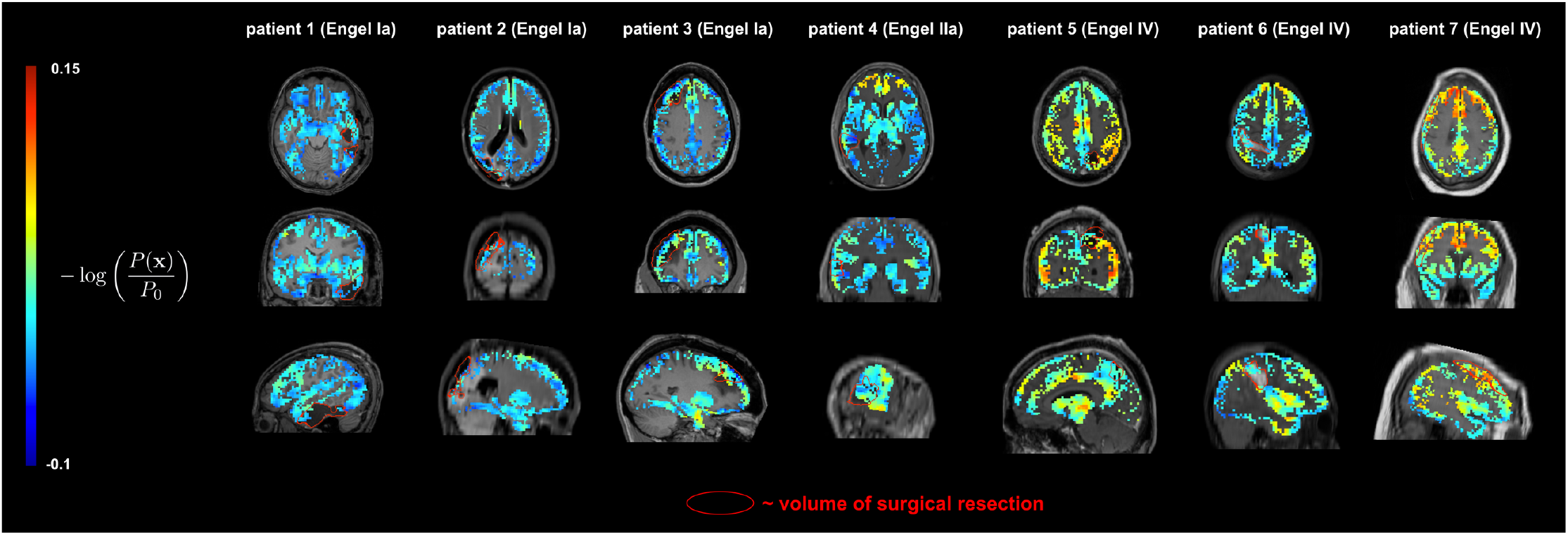
Describes reduced Gibbs distribution, P(**x**), for the absolute differences in functional connectivity between epilepsy patients and the averaged functional connectivity of 500 healthy control subjects drawn from the Genetics Superstruct Project, which is detailed in Figure 2. Compared to the Gibbs distribution of Figure 3, the reduced Gibbs distribution derives from correlations matrices that have been averaged within matrix columns. The resulting correlation vectors are ordered by unique spatial locations which are identifiable on a standardized atlas. Consequently, each element of a correlation vector describes correlation of BOLD time series from a spatial location with BOLD time series from other locations of the brain. Images from seven epilepsy patients indicate the information divergence, -log(P(**x**)/P_0_), of the functional connectivity of patients (in color), the boundary of surgical resections determined according to the standard-of-care (in red outline), and post-operative structural MRI (FLAIR or T1-weighted following contrast, gray scaled). Computation of probabilities used solely resting-state fMRI and structural MRI obtained before epilepsy surgery. Images from patients are ordered one column per patient with Engel Ia patients on the left, progressing to Engel IV patients on the right. Rows show transverse, coronal and sagittal views using radiological conventions.

Figure 5 shows patients, Engel classifications, and histograms of seed voxels for variant FC. Patients classified as Engel IV had histograms with broader distributions of FC with higher median score of variation of FC than patients classified as Engel Ia or IIa.

**Figure 5.**
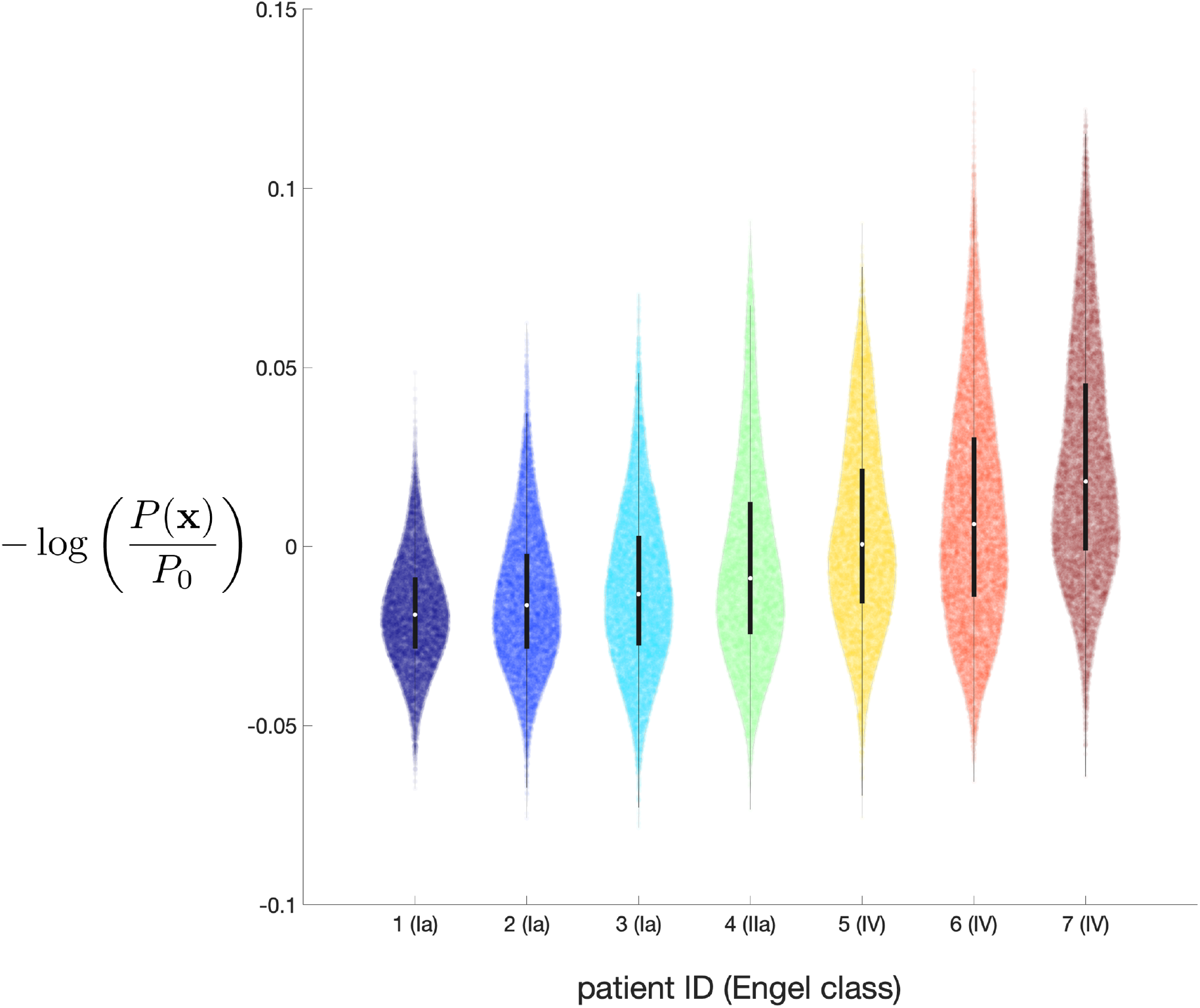
Describes reduced Gibbs distribution, P(**x**), for the absolute differences in functional connectivity between epilepsy patients and the averaged functional connectivity of 500 healthy control subjects drawn from the Genetics Superstruct Project, which is detailed in Figure 2. Compared to Figure 4, the information divergence, -log(P(**x**)/P_0_), is represented by violin histograms estimated from all anatomical voxels within gray-matter masks (18,611 voxels in 1000 histogramming bins). Embedded within each arbitrarily colored violin-shaped histogram is a black box plot indicating the median log probability of abnormality (circle) and central quartiles. Engel IV patients have median log probabilities > 0.

## Discussion

This study extends the method previously described by Stufflebeam et al. and is conceptually related to the method of Siegel et al. [11,12]. These prior works employed similarity measures, also based on correlations, to meaningfully compare blood-oxygen-level-dependent (BOLD) time series from a single patient with BOLD time series from a large cohort of control subjects. In this case series, we used related methodology to analyze rs-fMRI BOLD data from patients with refractory neocortical epilepsy who are candidates for epilepsy surgery.

Our findings support the hypothesis that epilepsy is a network disease with long-range and interhemispheric connectivity [25–28]. Work by Maccotta and colleagues using interictal rs-fMRI analysis in a mesial temporal lobe epilepsy cohort, with individually matched controls, also found that epilepsy exhibits characteristics of a network disease with patterns of variant FC involving widely distributed regions of the brain with organization by RSNs [29]. While the networks of mesial temporal lobe epilepsy could be fundamentally different than those of neocortical epilepsy, there is increasing evidence to supporting the hypothesis that neocortical epilepsy is also a network disease [30].

Work by Negishi and colleagues analyzed pre-operative fMRI using post-operative resection volumes as seed regions for analysis [26]. They noted that a focus on resection volumes in fMRI analysis may disproportionally emphasize short-range regional connectivity at the expense of long-range network effects in epilepsy. In our work, regions of highly variable FC did not precisely overlap with post-operative resection volumes. Most patients had regions of highly variable FC partially within or nearby resection volumes, but all additionally had hotspots in distant locations. The observed overlap between variant FC hotspots and resection volumes can also be affected by post-surgical brain morphology changes that influence the mapping of post-surgical volumes onto pre-surgical BOLD. Such discrepancies along with the presence of regionally diverse hotspots highlight the limitations of relying on post-operative resection volumes when analyzing FC both for structural and network related reasons. Furthermore, the absence of clear clinically determined SOZ boundaries may lead to standard of care resection volumes that, in retrospect and with evolving methods, may not be sufficiently aligned with the microscopically abnormal brain or regions that exhibit highly variable FC.

When looking at patients’ global trends of variant FC, we found that the three patients with Engel IV outcomes had the most variable and widespread FC when compared to a control cohort. These findings may indicate more diffuse disease in patients with Engel IV outcomes. This is particularly notable since clinical evaluation prior to surgery in the Engel IV patients did not suggest a predisposition towards poor post-surgical outcome. The pre-surgical evaluations of the Engel IV patients using traditional techniques were localizing just as the pre-surgical evaluations of Engel I and II patients. Work by Zijlmans et al. in 2007 used an EEG correlated fMRI technique to better stratify cases with complex SOZ localization as strong versus poor surgical candidates. This work used focal anatomic regions of variant BOLD as positive indicators of surgical candidacy, and did not offer SOZ resective epilepsy surgery to patients with non-focal BOLD abnormalities [31].

Patient seven did not undergo complete resection of their clinically determined SOZ due to the proximity of the SOZ to eloquent cortex, and instead underwent multiple subpial transections. This is meaningful as some long and short-range connections to the presumed SOZ likely remained post-operatively. It is estimated that only 16% of patients who undergo multiple subpial transections have long term seizure freedom post-operatively [7]. However, most patients (80%) do experience long term reduced seizure frequency [32]. This was not the case for patient seven who continued to have their baseline seizure frequency post-operatively. It is possible that this is related to more diffuse disease as suggested by highly variable FC results seen in this patient.

Two patients in this study, patients two and four (Engel Ia and IIa), had lesional neocortical epilepsy and five patients: 1, 3, 5, 6, 7 (Engel Ia, Ia, IV, IV, and IV), had non-lesionsal neocortical epilepsy. Epilepsy surgery in non-lesional neocortical refractory cases has a success rate of 34%. This is significantly less than the success rates for lesional neocortical and mesial temporal lobe epilepsy surgery, 66% and 69% respectively [6]. The difficulty in determining the location and margins of the SOZ make non-lesional refractory epilepsy particularly challenging to manage surgically. Currently, pre-operative non-invasive evaluation is used to identify a general region of the brain that is believed to contain the patient’s SOZ. Neurosurgical placement of iEEG then follows to better localize a SOZ for resection. These methods are limited by the area accessible by the electrodes via craniotomy (e.g. grid and strip electrodes) and by the number of electrodes that can be stereotactically placed. The limitations inherent in this work up can lead to incomplete characterization of SOZ and can contribute to confirmation biases [33]. Alongside further investigation and understanding of neocortical seizure networks, FC methods may contribute to localization strategies and resective approaches to epilepsy that identify and disrupt seizure networks. These methods can also contribute to the selection of patients most likely to benefit from surgery. It is therefore worthwhile expanding investigations of the use of rs-fMRI to investigate seizure networks that can be surgically treated.

Medical conditions in our patient cohort, including bipolar disorder, anxiety, migraines and hypothyroidism, may associate with abnormal FC. For example, personality traits that are predictors of depression and anxiety have been found to be more common among mesial temporal lobe epilepsy patients and have been associated with decreased frontotemporal synchronization on rs-fMRI analysis [34]. In our cohort, patients 5-7 had more underlying health concerns, had worse surgical outcomes, and had the most variable FC. The causal relationship between these comorbid conditions and abnormal epilepsy networks is unknown.

The use of rs-fMRI biomarkers as indicators of epileptogenic targets of surgery in medically refractory epilepsy has been suggested by work by Boerwinkle and colleagues, which found that network-targeted surgeries correlated with post-operative seizure freedom in a pediatric cohort [28]. One suggested mechanism for rs-fMRI’s utility as a biomarker of SOZs implicates increased energy demands of epileptiform activity, demonstrated by abnormalities in phosphocreatine to ATP ratios in mesial temporal lobe epilepsy [35]. Variations of FC in epilepsy patients may similarly have utility as a biomarker for interpreting disturbed BOLD signal in the context of medically refractory epilepsy.

Limitations of this study include the small number of patients, potentially confounding medical histories, and the study’s retrospective nature. Additionally, changes in brain contour within the skull following invasive work up and resective surgery introduce significant errors in coregistration of resection areas with pre-surgical rs-fMRI, creating challenges for associating pre-surgical rs-fMRI and the anatomy of surgical resections. Such errors in co-registration are difficult to resolve with current linear and nonlinear registration methods. Control subjects also originated from a different study and had fewer BOLD frames than study patients.

## Conclusion

The FC methods described in this work provide early support for the notion of neocortical epilepsy as a disease that results in regional RSN disruptions. In this retrospective case series, we found that patients with Engle IV outcomes had more variable and more widespread pre-surgical FC despite having localizing pre-surgical seizure scalp and intracranial EEG similar to their Engle I and II peers. One may hypothesize that these and similar imaging techniques will provide a valuable tool in pre-surgical planning if verified through larger and prospective works. Promising future work would investigate the possibility of seizure networks that can be disrupted. Such work would be particularly meaningful in cases of non-lesional neocortical epilepsy, where resection targets are difficult to delineate and epilepsy surgery outcomes are worse.

## Data Availability

All unidentifiable data, containing no protected health information, de-identified intermediate data, and software used in the present study are available upon reasonable request to the authors.

## Acknowledgements

JJL, KYP, JSS and ECL received funding from the NIH/NCI, R01 CA203861.

